# A qualitative process analysis of DCT as an alternative to self-isolation following close contact with a confirmed case of COVID-19

**DOI:** 10.1101/2022.01.14.21267257

**Authors:** Sarah Denford, Alex F Martin, Lauren Towler, Fiona Mowbray, Rosie Essery, Rachael Bloomer, Derren Ready, Nicola Love, Richard Amlôt, Isabel Oliver, G James Rubin, Lucy Yardley

**Affiliations:** Health Protection Research Unit in Behavioural Science and Evaluation, University of Bristol, Bristol, UK; Population Health Sciences, Bristol Medical School, University of Bristol, Bristol, UK; School of Psychological Science, University of Bristol, Bristol, UK; Health Protection Research Unit in Emergency Preparedness and Response at King’s College London, London, UK; School of Psychology, University of Southampton, Southampton, UK; UK Health Security Agency, England, UK

**Keywords:** Covid-19, lateral flow device, testing, process evaluation, qualitative

## Abstract

**Background:** In July 2021, a randomised controlled trial was conducted to compare the effect on SARS-CoV-2 transmission of seven days of daily contact testing (DCT) using lateral flow devise (LFT) and 2 PCR tests as an alternative to 10 days of standard self-isolation with 1 PCR, following close contact with a confirmed case of COVID-19. DCT appeared equivalent to self-isolation in terms of transmission in the trial, however it was not clear how tests were viewed and used in practice. In this qualitative study, we used a nested process to aid interpretation of the trial and provide insight into factors influencing use of tests, understanding of test results, and how tests were used to inform behavioural decisions.

**Methods:** Interviews were conducted with 60 participants (42 randomised to DCT and 18 randomised to self-isolation) who had been in close contact with a confirmed positive case of COVID-19 and had consented to take part in the trial.

**Results:** Sub-themes emerging from the data were organised into three overarching themes: (1) assessing the risks and benefits of DCT; (2) use of testing during the study period and (3) future use of testing. Attitudes toward DCT as an alternative to self-isolation, and behaviour during the testing period appeared to be informed by an assessment of the associated risks and benefits. Participants reported how important it was for them to avoid isolation, how necessary self-isolation was considered to be, and the ability of LFTs to detect infection. Behaviour during the testing period was modified to reduce risks and harms as much as possible. Testing was considered a potential compromise, reducing both risk of transmission and the negative impact of self-isolation and was highly regarded as a way to ‘return to new normal’.

**Conclusion:** Participants in this study viewed DCT as a sensible, feasible and welcome means of avoiding unnecessary self-isolation. Although negative LFTs provided reassurance, most people still restricted their activity as recommended. DCT was also highly valued by those in vulnerable households as a means of providing reassurance of the absence of infection, and as an important means of detecting infection and prompting self-isolation when necessary.

## Background

Use of lateral flow device (LFT) antigen tests to provide a rapid diagnosis of COVID-19 infection has been an integral part of the UK Government response to the pandemic. In the Autumn of 2020, mass testing pilot schemes were introduced so that members of the public could take a test even if they did not have symptoms of COVID-19 [1, 2]. By April 2021, members of the public were encouraged to get tested with a LFT twice weekly [3, 4] so that asymptomatic and pre-symptomatic infections could be identified. Those testing positive for COVID-19 were required to take a confirmatory PCR test and self-isolate for 10 days. Close contacts of the positive case were also required to self-isolate at home for 10 days. Although self-isolation is an effective strategy for reducing the spread of COVID-19, it can have a substantial and negative impact on the individual and society [5, 6]. Self-isolation following close contact remains a requirement for adult contacts who are not fully vaccinated.

In an attempt to reduce the negative impact of self-isolation without increasing transmission of COVID-19, the government made available a scheme in which daily contact testing (DCT) with an LFT was offered as an alternative to self-isolation for some settings [7-9]. Previous research has suggested DCT may be a feasible alternative to self-isolation [10-13], and may be as effective as self-isolation for controlling transmission in certain situations [14]. However, earlier studies were either conducted while stringent society-wide restrictions were in force, and familiarity with testing was low [11, 12], or within specific settings, such as schools [14]. Research is needed to understand how the general public view and use testing as an alternative to self-isolation in a context in which many government imposed restrictions have eased and familiarity with testing has increased.

In July 2021, Public Health England (now UK Health Security Agency), supported by researchers at the University of Bristol and King’s College London (KCL), conducted a randomised controlled trial of the impact of DCT for contacts of COVID-19 cases on transmission in the UK. The primary aim was to compare the effect on infection transmission of seven days of DCT as an alternative to 10 days of standard self-isolation following close contact with a confirmed case of COVID-19. Participants randomised to DCT (DCT group) had the option to take a lateral flow test for seven consecutive days and were granted freedom from self-isolation for a 24hour period on receipt of a negative test result.

Participants in the DCT arm were also asked to take two PCR tests; one on performing their first LFD and one after testing positive/on last negative LFD. Those in the standard self-isolation (SI group) were asked to take a single PCR test and self-isolate for 10 days.

A total of 49,623 close contacts took part in the trial (excluding ineligible and withdrawing participants), of which 4,006 participants submitted a positive PCR test result. The trial found that the proportion of secondary cases from contacts of those who were in the DCT group (6%) were comparable to those who self-isolated (7.5%) [15]. The trial also found similar results among people who had received at least one dose of the vaccination (6.9% in the DCT arm compared with 7.8% in the SI arm).

While a policy of DCT appears equivalent to self-isolation in terms of the risk of onwards transmission, previous research has highlighted a range of concerns and uncertainties that members of the public have, from concerns about the accuracy of LFT to confusion about how to engage with the policy if a household member has COVID-19 [11]. These issues may affect how people use LFTs and limit their willingness to use the freedoms that the system allows. Therefore, a secondary aim of the trial was to use a mixed methods approach to examine the acceptability and feasibility of DCT as an alternative to self-isolation, and behaviour during the testing period. This aim was achieved by means of a survey of 20,004 (40% response rate) participants who completed the trial, and in-depth one to one interviews. The survey analysis (reported elsewhere [15]) compared those who were DCT and only reported negative tests (DCT negative test) to those who reported at least one positive test (DCT positive test) and those who completed standard self-isolation (SI), and found that most participants, regardless of group allocation, reported modifying their behaviour during the study period. Among those who were supposed to be isolating (i.e., those in the In the SI and the DCT positive test group) approximately 4 out of 5 people reported much less contact with non-household contacts during the study period. Among those who were not required to isolate, 3 out of 5 people reported much less contact. Only a small number of participants reported that they had left the home whilst self-isolating; approximately 1 in 6 in each group. However, the most common reason for leaving the home whilst isolating was to take a COVID-19 test. The survey also indicated that participants were confident in the results of their tests, with 79% of SI participants reporting that they were very or completely confident in the accuracy of their PCR test results, 64% of participants in the DCT positive group (PCR and LFT) and 83% in the DCT negative group.

In this qualitative study, we used a nested process to aid interpretation of the trial and survey findings by providing a detailed understanding of factors influencing the use of tests, understanding of test results, and how tests are used to inform behavioural decisions.

## Methods

### Design

We conducted interviews with individuals who had been in close contact with a confirmed positive case of COVID-19 and had consented to take part in the trial of LFT as an alternative to self-isolation.

### Sampling and data collection

At the time of recruitment to the trial, all participants were asked if they would be happy to be contacted by researchers from the University of Bristol and King’s College London. Demographic and contact details of those consenting to contact were shared securely with the research team. We carried out a pilot study prior to the RCT, findings have been reported previously [10, 11]. Data collected as part of the pilot study suggested key factors that may influence acceptability of DCT, and purposive sampling was used to ensure diversity in those factors, including; trial group allocation, gender, ethnicity, date of initial contact, and whether the participant lived in the same household as the confirmed positive case. Selected participants were contacted by text, phone, or email, and provided with a study information sheet. All interviews were conducted once the participant had completed the period of testing or self-isolation, and participants were given a £40 shopping voucher as reimbursement for their time.

Interviews were conducted remotely (online or by telephone) by a qualitative researcher (SD, AFM, FM, LT, GT, BA, RAE, and RB) between the 24^th^ June and the 8^th^ July 2021. Our initial topic guide was based on findings from a qualitative analysis of the related pilot study [11] and designed to include open questions to explore experiences of the testing process, beliefs about testing, perceptions of positive and negative test results, and the impact of testing on behaviour. In order to encourage participants to speak openly about their views and behaviour during the testing period, participants were informed that the interviews would remain anonymous even if they disclosed having not always adhered to the guidance. However, participants were reminded that the research team would be obliged to notify authorities if the participant revealed any serious intended or planned breaches of COVID-19 regulations that could put others in danger. In practice, we did not need to make any notifications.

All participants provided verbal consent prior to taking part in the interview. Ethical approval was granted by the Public Health England Research Ethics and Governance Group (Reference NR0235).

### Analysis

In accordance with the stages of thematic analysis [16] anonymised transcripts were read by two authors (SD, LT) and detailed notes were made about interesting concepts and ideas. Using the software NVivo 12, all text was labelled with an initial set of codes. Through discussion, similar codes were combined, and a preliminary set of themes were agreed. Relevant data for each theme were collated and reviewed, and themes refined and defined [16].

### Results

A total of 60 participants took part in an interview, including 42 (70%) participants randomized to DCT (DCT group), and 18 (30%) who were randomized to 10 days self-isolation with a single PCR (SI group). Of those randomised to the DCT group, 18 (43%) lived in the same household as the positive contact (DCT household positive group). Of the total participants, 33 (55%) were women, and 30 (50%) were from an ethnic minority background.

## Results of the thematic analysis

### Assessing the risks and benefits of DCT

Attitudes toward DCT as an alternative to isolation appeared to be informed by an assessment of the associated risks and benefits. Participants considered how important it was for them to avoid isolation, how necessary self-isolation was, and the ability of LFTs to accurately detect infection. Testing was considered a potential compromise, reducing both risk of transmission and the negative impact of self-isolation, and was highly regarded as a way to ‘return to new normal’.

### Importance of avoiding isolation

Participants varied in the extent to which they wanted or needed to avoid isolation. Participants discussed multiple negative implications of self-isolation on mental and physical health, as well as the impact on their ability to work and receive an income. Those who were positive about the use of DCT explained how testing had the potential to reduce the negative impact of isolation:

*“I think [testing] prevents isolation, I think for myself being stuck indoors is really bad for your mental health and your physical health, so I think for me the biggest thing was being able to kind of go out and get some fresh air” (DCT 013)*.

*“Definitely, I’d rather [test] than isolate yeah. Absolutely…We don’t have anything to cover us if we are out of work we just don’t get paid that’s it” (DCT 007)*.

Multiple lockdowns and isolation periods increased the extent to which people were motivated to avoid additional restrictions, and testing was often viewed as a potential lifeline:

*“There’s a limit to how many times you can do [self-isolation] and still be employed” (DCT 015)*.

*“It gives you the potential that you can go out, and I think now we’ve spent so much time inside I think it’s quite important that if we can get out then we should, so yeah, definitely” (DCT 001)*

Perceived benefits of testing were lower among those who were able to work remotely, had a supportive network, or were happy to spend time alone:

*“Things like food and stuff weren’t an issue cos obviously my husband could still go out and get stuff. I’m not massively fussed about going out gallivanting or anything like that, so I’m more than happy in my own company for a few days” (SI 020)*.

### Perceived need for self-isolation

Attitudes toward testing were strongly influenced by how necessary the participant considered self-isolation to be. Many participants did not consider self-isolation to be necessary because they did not consider themselves likely to have caught the virus. This may have been because they considered the contact with the positive case to have been low risk; for example, contact had not been prolonged, inside, or they had not been in close proximity with the confirmed case:

*“The person that was close-contact with – I would have said I was always two metres away from her… in that garden… I thought I’d kept me distance from her, but Track and Trace said because we were in the garden, I would still be classed as a close contact” (SI 003)*.

*“No, [I did not think I had caught COVID-19] not at all, no. That’s what really annoyed me*… *when my daughter said, ‘Dad, you’ve got to start doing the ten-day stay-in now, isolation*.*’ I said, ‘what you on about?’ She said, ‘well, where you’ve walked in that house, they reckon if you’ve walked in a house where they are infected you can get infected’” (DCT 012)*.

Other participants did not consider themselves likely to have caught the virus because of their infection control measures, or because they were fully vaccinated:

*“I’d obviously had both my vaccinations as well so I thought the possibility of me then catching COVID was pretty minimal” (DCT 004)*.

*“No, I don’t think [I had caught the virus], because I didn’t have any symptoms… I’m fully vaccinated now as well” (DCT 002)*.

A low perceived likelihood of infection often led to frustration when participants were told that they may have to self-isolate, and increased acceptance of DCT:

*“I can’t believe I need to do [isolation] again because I’ve been in contact with somebody, I don’t have COVID” (DCT 014)*

*“I thought [DCT] was a very sensible move – I felt very happy about it… because self-isolation just seems really unnecessary and a bit extreme, in my opinion, whereas this being able to do the test and then leave the house… I just think that is a sensible step forward… I’m testing negative… [isolation] just seems unnecessary” (SI 004)*.

Likewise, those who had been vaccinated described how this positively influenced attitudes toward testing:

*“I think it’s going to be a progression in conjunction with the vaccine, and what was proposed during the trial seemed to be very much the common sense solution when you know individuals have been vaccinated, or when there’s been shall we say questionable or short term contact with individuals [testing] seemed like a prudent step to take” (DCT 003)*.

Perceived likelihood of infection was higher among those who viewed themselves as having been in close contact, or were living with, a positive case. This group voiced greater concerns about the safety of DCT, and at times, considered isolation to be a better option:

*“I, obviously, live with my daughter and she’s a child, so we have loads of close contact, so I thought I would get it. I was quite concerned about that” (DCT household positive 047)*

*“But I think it would probably depend on the circumstances in which I’d come in contact with this person. If it was another person my household again like it was in this circumstance, I’d probably be inclined to stay indoors again for most or all of the isolation period. If however I’d been notified through the COVID app that I’d been in contact with someone for five ten minutes I might be more likely to go, ‘Okay well the chances I caught COVID off that person are probably fairly slim’, I’d feel more comfortable doing DCT, going out and about and hopefully not putting anyone else at risk” (DCT household positive 039)*.

### Accuracy of test results

Use of DCT as an alternative to self-isolation appeared to be influenced by participants’ understanding of the accuracy of LFTs. Participants seemed divided as to how well LFTs were able to detect the virus. Whilst some participants were confident that the test would be as accurate as it could be, others were less certain:

*“I am aware that lateral flow can produce false positives, but it’d be in the 90%, 95% to 99% [accurate] I would have said - I don’t know what the actual figure is of accuracy, but that’s where my imagination lies” (DCT 023)*.

*“I knew it was basically it was going to be at least 90 to 100% it was going to be inaccurate” (DCT household positive 026)*.

Confidence appeared to be reduced by experience of conflicting test results, or participants holding conflicting beliefs regarding whether they were infected or not:

*“I don’t really have much confidence in the lateral flow tests. I don’t think they’re very accurate. For example, when my partner tested positive and we were isolating, he had a positive PCR and then like a day later took the lateral flow just to see and it said negative, but he did very much have COVID, so I don’t really have that much confidence in them” (SI 018)*.

*“[testing] almost feels pointless to be honest cos yeah I’ve never tested positive and there’s no way I’ve not had it” (DCT 007)*.

However, whilst it was accepted that tests were not infallible, they were often considered to be preferable to no testing:

*“It’s like any test – I’m sure you can’t say it’s hundred per cent accurate – there is always going to be some instances where it’s not giving you the right result, but I’m not really concerned. It’s the best thing to go by – what else would you do?” (SI 004)*.

Concerns about the accuracy of the tests could lead to concerns about the safety of permitting people to leave isolation based on a negative lateral flow test result:

*“Personally I thought a bit, oh what’s the word I’m looking for? Unclean because even though it was negative and even though I had like I say it was legally now allowed to leave the house, I still knew that it wasn’t a 100% and I could still possibly infect somebody else” (DCT household positive 026)*.

Participants, particularly those who felt they needed to avoid self-isolation, often described attempts to increase accuracy of testing, for example, through using multiple tests, or using tests in combination with other infection control measures to maximise safety:

*“[I went] to the chemist, got some lateral flows and I did one, yeah it came back straight away as negative. I did a second one just to make sure…I wanted two to compare, so the chances of both of them being a false negative was kind of remote …” (DCT household positive 026)*.

### Exposure to vulnerable groups

Regardless of perceived likelihood of having the virus, and beliefs about the accuracy of tests, the majority of participants did not feel confident having contact with vulnerable people:

*“I think the only thing probably I wouldn’t have felt 100% comfortable doing is going into the office because obviously where I work there are some people that would class themselves as older or vulnerable” (DCT 004)*.

*“There is no way [my wife would] have been able to go into a hospital where you’ve got vulnerable children… so I think there should be a different set of guidelines, so it almost ratchets what you can do up or down” (SI 010)*.

### Use of tests during the study period

Knowledge of how tests should be used during the study period

Participants varied in their reports of information they had received regarding the rules and regulations for DCT, and although some participants thought the rules were very clear, others disagreed:

*“It might just be me, but it just seemed like there wasn’t, it wasn’t as clear… are you allowed to do BAU [business as usual] or, is more the idea of you doing just the actual minimal activities, what’s the difference? Are you expected to live your best life for the ten days or whatever or is the idea more, look you’re allowed to leave but be as minimalistic as you can? I think that’s where -- and I couldn’t answer that question because I didn’t know” (DCT household positive 041)*.

It was thought that greater clarity could be helpful, as some participants reported having to seek the information out themselves:

*“I guess that would be the thing that I would say about this, is that it would be useful to have a bit more definition around what can and can’t be done…. it would help to have just a bit more definition around what is and isn’t essential… I didn’t find it super-clear in terms of what you were and weren’t allowed to do” (DCT 016)*.

*“I had to Google it myself and find out because yeah, I didn’t get sent anything at all that I couldn’t do, it just said this means you don’t have to self-isolate, it didn’t say anything else than that” (DCT household positive 002)*.

### Facilitate engagement in low-risk activities

Many participants in the DCT group described how testing had enabled them to engage in low-risk and essential activities such as exercise, shopping, and to collect prescriptions during the testing period:

*“So I was able to go to the shops and help with the pick-up and drop off of the kids, and just able to go out for exercise. I’d go to the park and also take the kids to the park” (DCT 004)*.

Among those who considered themselves to be at high-risk of having caught the virus, daily tests provided an additional layer of reassurance that they were safe to engage in low-risk activities outside the home without transmitting the virus to others:

“*I wouldn’t want to go and infect people, and I would prefer the peace of mind to know that I’ve done a test and it’s negative. It would just make me feel more confident that I was okay to go out and about” (DCT household positive 044)*.

However, this group also described efforts to minimise close contact as much as possible; either through choosing to go out at quieter times of the day, or using infection control measures to reduce risk of transmission to others:

*“Yes, I minimised contact. Didn’t see anybody, anybody that I’m familiar with. Didn’t do shopping. Didn’t do any crowded spaces. Honestly, all I did was my normal kind of walking or running” (DCT household positive 041)*.

*“We just kept to ourselves and kept our distance, and went out later when there was fewer people about anyway, so it was almost a bit of a throwback to the start of the pandemic” (DCT household positive 043)*.

Those considering themselves to be unlikely to have caught the virus still described feeling reassured by test results, describing the role of testing in reducing any remaining element of doubt:

*“It’s reassurance for us doing it, or me doing it, and people I’m people I’m around, even though I wouldn’t tell them I was doing it, because I don’t believe I’ve ever had [COVID-19], but in my head I’ll know that I wouldn’t be a risk to them” (DCT household positive 043)*.

Even those who were concerned about the accuracy of tests were able to use tests as an additional layer of reassurance prior to leaving the home:

*“I’d like to have the option [of testing] I think, yeah*… *I don’t have, as mentioned, complete confidence in the lateral flow tests, they did test positive I presume for most symptomatic points of my – of my housemates experience of COVID and so that lined up with the results coming from the PCR test so I feel as though they were accurate for two or three days of her illness which were probably the points at which she was most infectious or posing a risk to others so in that seen it would have been useful to know yeah if I was infections and if I was posing a risk to others” (DCT household positive 039)*.

However, as with those who considered themselves to be at high-risk of having caught COVID-19, those with concerns about the accuracy of tests also reported employing extra caution during the testing period:

*“I was permitted to go out and have exercise and visit essential shops… I very much was not going to trust that negative result as, you know, a cart blanche to go out and assume I didn’t have coronavirus, I think I still treated it with a degree of caution having seen that my you know symptomatic and then tested positive housemate had been returning negative lateral flow tests so I think I approached it with relief but caution” (DCT household positive 039)*.

### Provide reassurance and peace of mind

Participants in both the DCT and the self-isolation/PCR group who considered themselves likely to catch, or have caught, the virus described using testing to reassure themselves and their housemates that they were not infected:

*“I’d rather do [a test] anyway just to make sure you’re safe” (DCT 012)*.

For those living with a positive case, and so in constant contact with the virus, the tests provided regular reassurance that they had not caught the virus:

*“The way I understand it is if you’re living with a person who has it, that person’s contagious for ten days. So there was a bigger concern that, theoretically, on day seven to whatever near the end of the quarantine period we could get infected. So, by taking a test every day it actually gave me that reassurance that I wasn’t infected if that makes sense. So actually did the opposite of what you’re asking. The tests had reassured me that I wasn’t getting infected daily, if that makes sense” (DCT household positive 041)*.

*“It was good peace of mind to know that, even though someone in the house had COVID, I was still testing negative” (DCT household positive 044)*.

Even those who had been fully vaccinated, and as a result considered themselves unlikely to have contracted COVID-19, reported feeling safer both at home and outside the home as a result of DCT:

*“The fact that I was testing and I knew that I’d had two jabs, yeah it made me feel a lot safer” (DCT household positive 012)*.

Participants in the self-isolation/PCR arm described using LFTs during the study period for reassurance purposes, particularly if they considered themselves or their household to be at risk from COVID-19:

*“I have a lot of lateral flow tests at home, so I did a test immediately and tested negative thankfully. Then I tested myself every day for five days and tested negative” (SI 025)*.

*“‘Because she’s also high-risk because she’s got asthma, so we were just like, ‘We just need to keep testing and if anything changes, tell each other immediately’” (SI 046)*.

### Future use of testing

Testing was often considered to be a way out of the pandemic and a compromise between the need to avoid isolation and keep others safe:

*“I don’t see that as a big price to pay, really, for being able to go out, but also making sure everyone else is safe, and I can’t really think of another way to make it easier without perhaps increasing the risk factor for somebody else. It’s safe to go out and it allows you to have your life back” (DCT household positive 043)*.

*“We’re not potentially locking down or isolating people who haven’t got it and are never likely to develop it, but yet we’re still protecting people” (DCT household positive 043)*.

It was suggested that a policy of self-isolation for all contacts was unsustainable long-term, and testing was viewed as potential step toward normality:

*“Well, we can’t live like this for ever - half the country would be in isolation at any one time – so I was aware that this was obviously a first step towards a middle way of, actually, people could not continue the country with people isolating unnecessarily” (DCT household positive 043)*.

Those who had been double vaccinated felt that they could safely avoid self-isolation following close contact with a positive case if they also had a negative lateral flow test. However, even those who were double vaccinated appreciated having the option to take a test:

*“If you’ve had the vaccine there needs to be a bit of freedom now, because otherwise people are just going to do it anyway because they’re fed-up. So we just need to get back to a bit of normality now” (DCT 018)*.

*“I was very pleasantly surprised that there was a possibility for me to go outside especially as I’m vaccinated and know that I’m less likely to be infectious and that I was regularly taking lateral flows” (SI 025)*.

Indeed, participants frequently described a preference for testing should mandatory self-isolation for contacts be removed, often considering testing a small inconvenience for increasing safety and reducing transmission:

*“It does open up that flexibility that you can go places and do things if you need to or you want to. But at the same time, I wouldn’t want to do it without doing those tests because that’s sort of reassuring and okay” (SI 006)*.

It was thought that this had the potential to facilitate adherence to self-isolation through providing confirmation of infection:

*“I think if I tested positive, and I knew that I was a threat to other people – perhaps even if I’m not ill myself – then I can justify having to stay at home” (SI 004)*.

## Discussion

Given that infection transmission and self-isolation both have a major impact on health and society, it is essential to explore ways in which to reduce both transmission and the negative impact of self-isolation. A large trial across England found DCT to be a safe alternative to self-isolation facilitating greater return to normality and enabling people to carry on with work and other essential activities while controlling transmission [15]. This nested qualitative study found that, for most participants, DCT was viewed as a sensible, feasible, and welcome means of avoiding unnecessary self-isolation. This view was more commonly expressed by those who viewed their situation as low risk (for example due to their household being fully vaccinated, or the level of contact with the positive case being limited). DCT was also highly valued as a means of providing reassurance of the absence of infection, and as an important means of detecting infection and prompting self-isolation when necessary. This view was more commonly expressed by people who believed they had a high-risk of being infected, or who were concerned about serious consequences of infecting vulnerable people (for example, at home or at work). While there was some evidence that negative LFTs were reassuring for people, most people still restricted their activity as recommended. Participants expressed some uncertainty and confusion about the rules regarding what was and was not permitted during the trial and requested some clarity around what constitutes “essential” activity.

Despite concerns that DCT could increase contact and transmission, this was not found to be the case [15]. Survey data suggests that many participants in the trial modified their behaviour during the study period, and the current qualitative study may present some additional insight into this decision-making process. In accordance with survey findings, participants in the current study described how testing had facilitated engagement in low-risk activities. Behaviour during the testing period appeared to be the result of a carefully considered assessment of the risks and benefits of DCT; including the need to avoid isolation, the perceived likelihood of infection, and the accuracy of LFTs [11, 12]. In line with previous studies [11, 12], those considering themselves at a higher risk of having caught COVID-19 reported considerable efforts to reduce risk, with many avoiding close contact with others even after receiving negative test results.

Survey data collected alongside the main trial revealed that most participants were confident that tests were accurate, although those who used daily LFTs and reported a positive test had lower confidence compared to other participants. Data collected through the interviews reported here showed that participants were aware that LFTs did not always capture infection. Tests were often interpreted in combination with other indicators, such as presence or absence of symptoms, likelihood of contact with the positive case, or vaccination status. In some cases, participants attempted to increase accuracy through repeated use of LFT or PCR tests. However, whilst LFTs were often viewed as preferable to no tests, it was noted by participants that they should be used cautiously in combination with reduced contact and increased infection control behaviours as much as possible.

As of 16^th^ August 2021, in the UK only unvaccinated populations are required to isolate following close contact with a positive case, the trial found that the proportion of secondary cases were similar among DCT participants and IS participants who were double vaccinated (7.5%) [15]. Importantly, the 7.5% comprised mainly household contacts and very few non-household contacts were reported. Data generated through interviews also suggests that many participants were making educated assessments about the presence or absence of infection based on vaccination status (or degree of contact with infected cases). Even among the participants who were confident that they were unlikely to have contracted COVID-19, tests were able to provide an additional level of reassurance that they were safe to leave the house to engage in low-risk activities. This suggests that the option to take a daily test following close contact with a positive case may be welcomed by some members of the population regardless of vaccination status.

The current study also provides support for the role of DCT for providing reassurance to vulnerable populations as we move into the next phase of the pandemic. Indeed, participants in both the self-isolation and testing arm reported using LFTs throughout the study period to provide an early indication of the presence or absence of infection. Testing for reassurance appeared to be particularly important for those living in vulnerable households, who were able to use DCT alongside infection control measures to try to keep vulnerable members of the household protected from the virus. Those receiving a positive test result could isolate accordingly.

Previous studies have found DCT to be an acceptable alternative to self-isolation during lockdown [10, 11] and in school settings [12]. Interviews conducted as part of the current study occurred during the summer months with relatively few social distancing measures in place; opportunities for social interaction were relatively high, and the number of fully vaccinated people had increased. Whilst this rapidly changing context will inevitably shift the weight of perceived risks and benefits, participants still appeared to view DCT as an acceptable alternative to self-isolation, often considering DCT to be a potential compromise that could reduce the risk of transmission and reduce the impact of unnecessary isolation.

### Limitations of this work

Despite our best efforts to recruit a diverse sample of participants, the main potential limitation of this work is that relevant voices may have been missed. It is possible that more vulnerable populations, or those with greater concerns about the role of DCT as an alternative to self-isolation did not consent to take part in the trial and were therefore not invited to take part in an interview. It should also be noted that this work was conducted during a period when cases of COVID-19 were declining and restrictions were easing. This may have influenced perceptions of testing and isolation, and our results should be interpreted with this in mind.

### Implications for policy

This work has a number of implications regarding the use of testing in the future. As we negotiate a return to a ‘new normal’ it is essential that strategies and policies are introduced that maximise safety whilst reducing the negative impact of unnecessary isolation. DCT as an alternative to self-isolation has the potential to reduce the negative impact of self-isolation among those who are still required to do so. However, it may also provide additional reassurance, both inside and outside the home, for those who are not. At the time of writing, LFTs were freely available to members of the public in the UK, but this is not guaranteed to continue [17]. Our results suggest that there may be value in continuing to make tests freely available to people who have been in contact with a COVID-19 case, both to prompt early self-isolation when necessary and also to provide people with the reassurance needed to continue with their day to day activities.

### Conclusions

Participants in this study viewed DCT as a sensible, feasible, and welcome means of avoiding unnecessary self-isolation. Although negative LFTs provided reassurance, most people still restricted their activity as recommended. DCT was also highly valued by those in vulnerable households as a means of providing reassurance of the absence of infection, and as an important means of detecting infection and prompting timely self-isolation when indicated. Greater clarity around what constitutes essential activities would be welcomed.

## Data Availability

No additional data is available

## Declarations

### Ethics approval and consent to participate

Ethical approval for this study was granted by Public Health England’s Research Ethics and Governance Group (ref R&D 434). All participants provided informed consent/assent to participate. Informed consent was also obtained from parents of all participants under the age of 16 years. All methods were carried out in accordance with relevant guidelines and regulations.

### Consent for publication

NA

### Availability of data and materials

The datasets used and/or analysed during the current study are available from the corresponding author on reasonable request.

### Authors’ contributions

Conceived the study: All authors

Study design: All authors

Analysed the data: SD

Interpreted the data: All authors

Drafted the manuscript: SD

Reviewed the manuscript and approved content: All authors

Met authorship criteria: All authors

## Acknowledgements

LY is an NIHR Senior Investigator and her research programme is partly supported by NIHR Applied Research Collaboration (ARC)-West, NIHR Health Protection Research Unit (HPRU) for Behavioural Science and Evaluation, and the NIHR Southampton Biomedical Research Centre (BRC).

SD, RA, RAE, and IO are funded by the NIHR Health Protection Research Unit (HPRU) in Behavioural Science and Evaluation at the University of Bristol.

GJR and RA are also funded by the NIHR Health Protection Research Unit (HPRU) in Emergency Preparedness and Response at King’s College London.

AFM is supported by the Economic and Social Research Council Grant Number ES/J500057/1

---

**Table 1:**
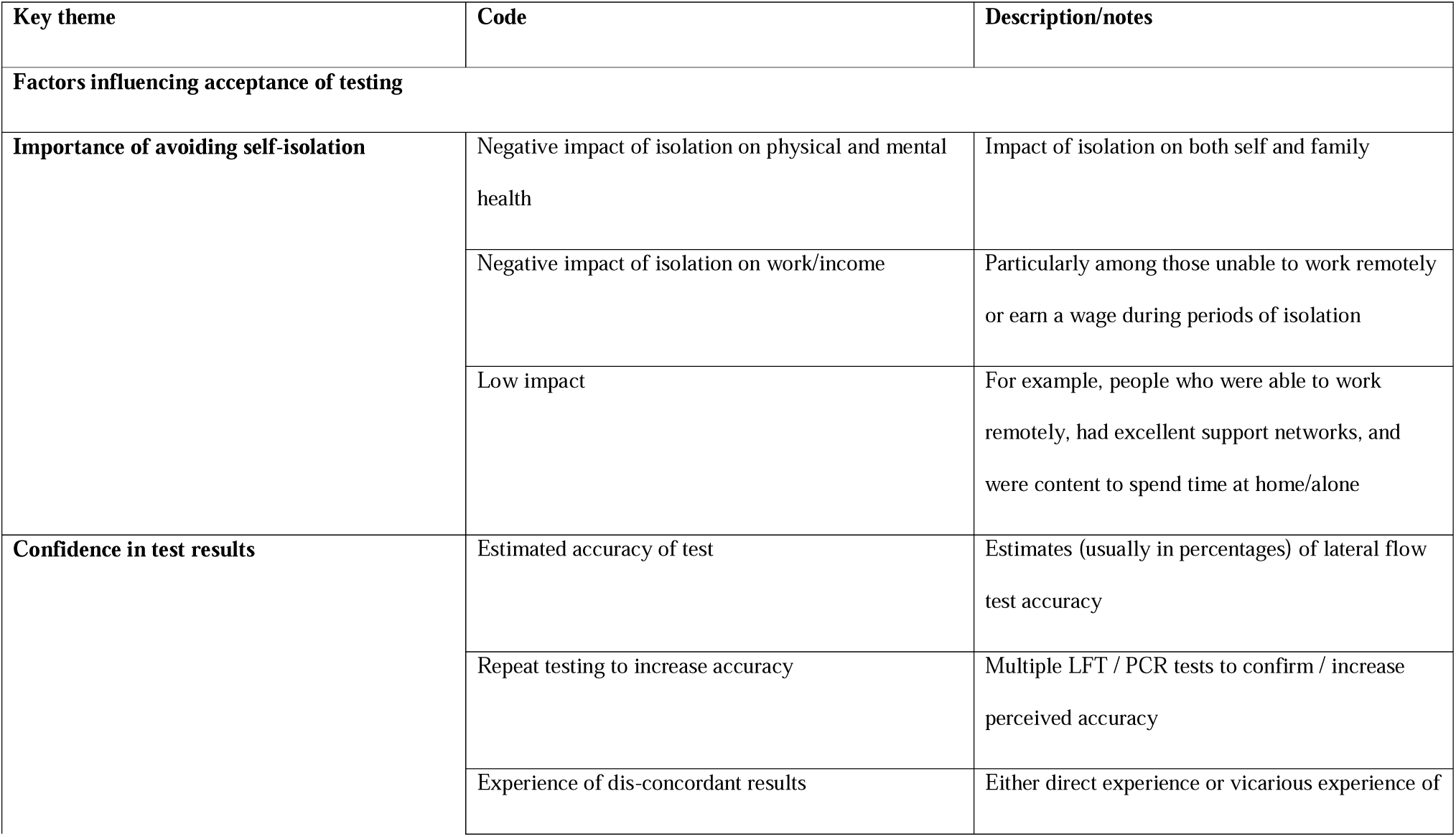
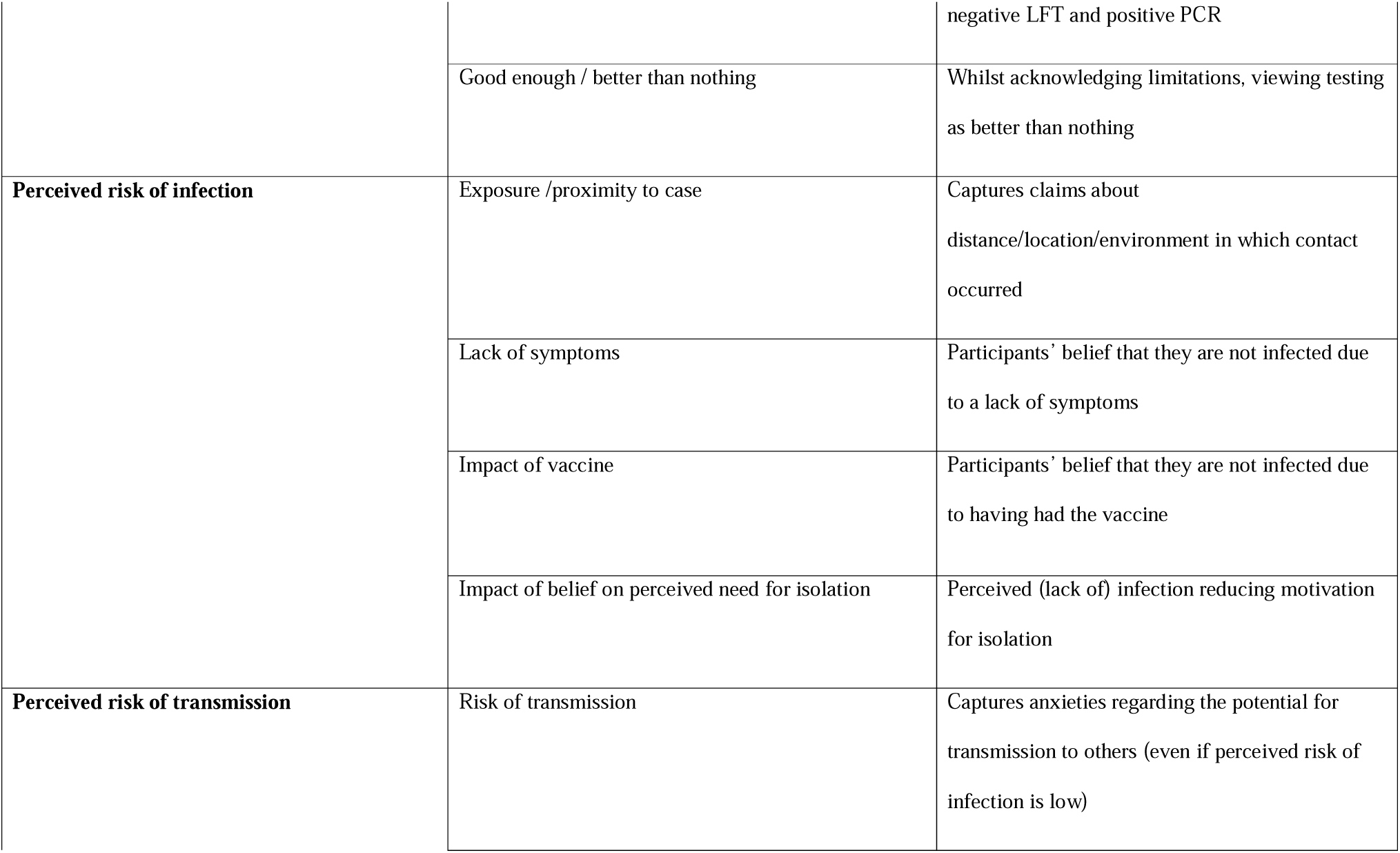
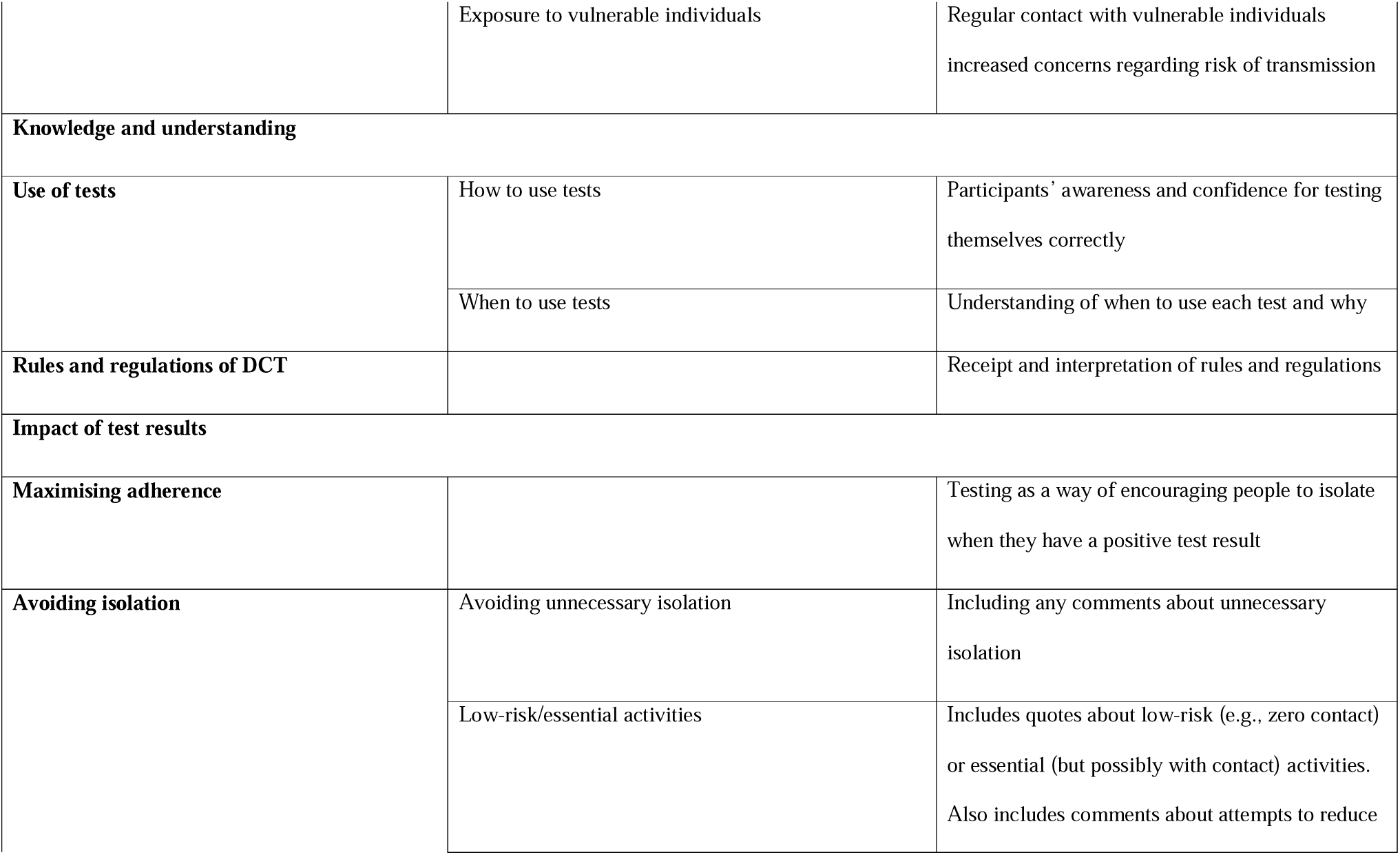
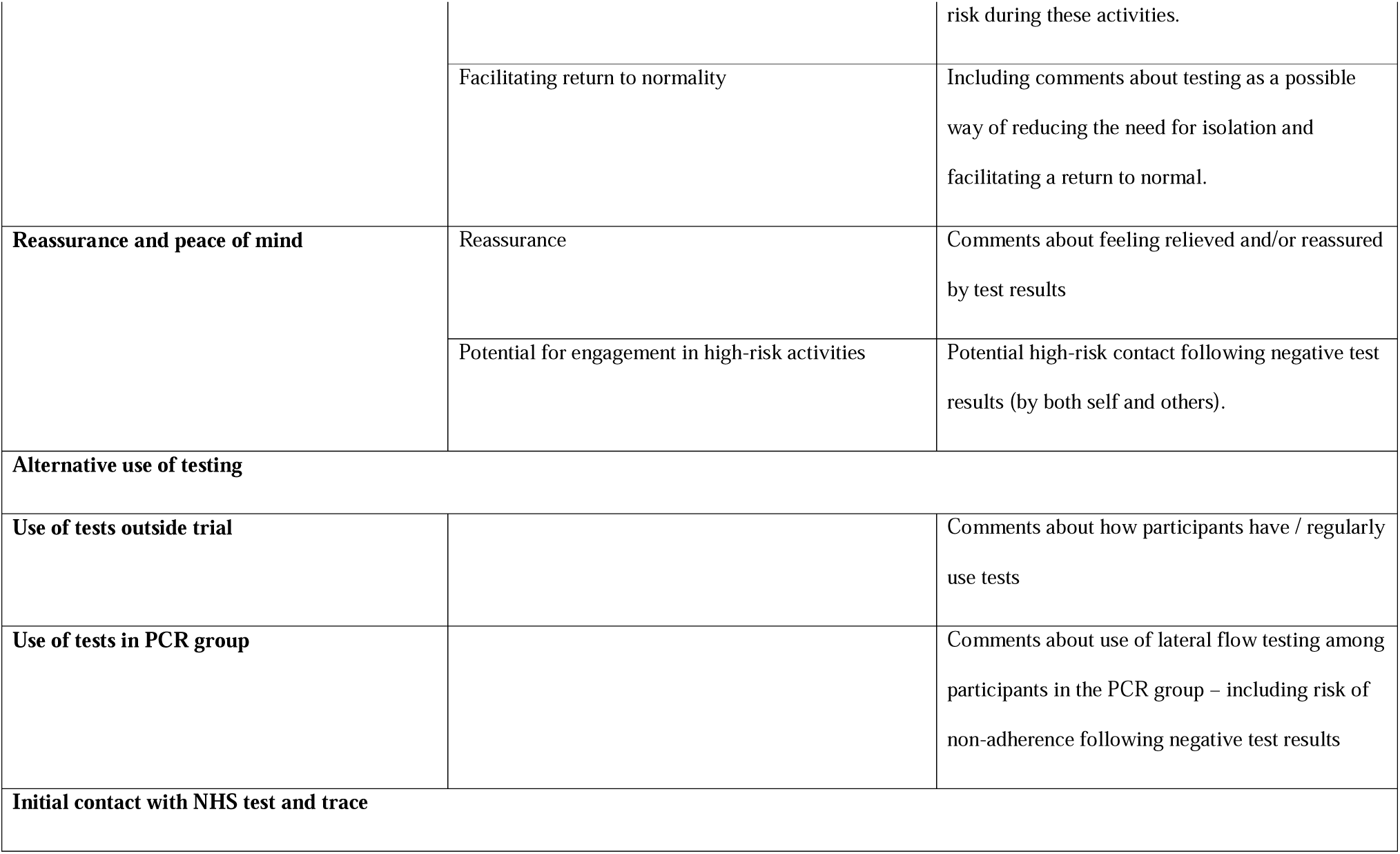
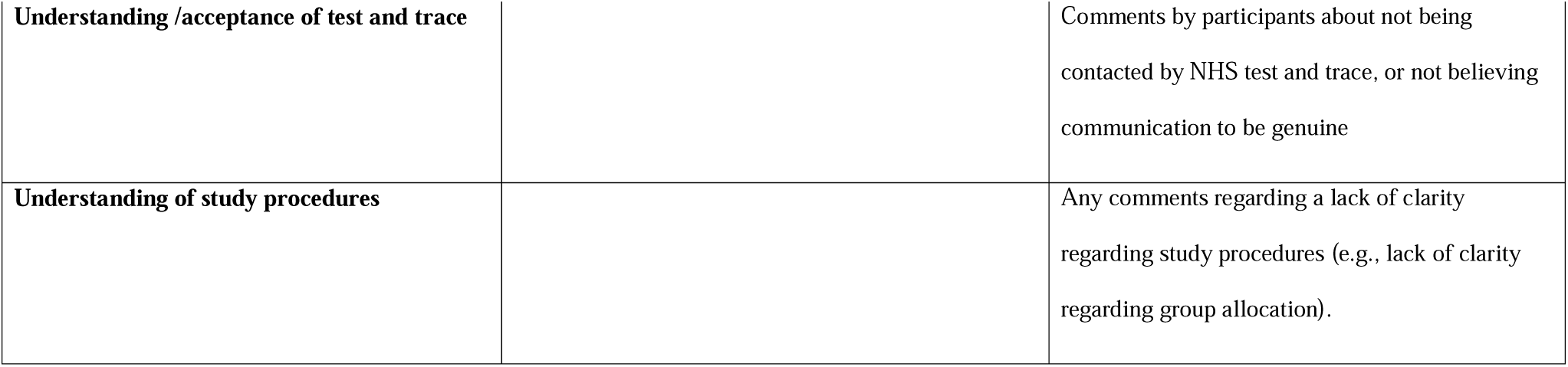
Initial coding framework

